# Cost-effectiveness of direct oral anticoagulant management programs

**DOI:** 10.64898/2026.03.12.26348295

**Authors:** Jordan B. King, Catherine G. Derington, Stanley Xu, Nathan P. Clark, Kristi Reynolds, Jaejin An, Daniel M. Witt, Maureen O’Keeffe Rosetti, Daniel T. Lang, P. Michael Ho, Elissa M. Ozanne, Brandon K. Bellows

**Author notes:** **Address for correspondence:** Jordan King, PharmD, MS; 295 Chipeta Way, Williams Building; Salt Lake City UT 84108.

## Abstract

**Background:** Pharmacist-led anticoagulation management services (AMS) for direct oral anticoagulants (DOACs) reduce prescribing errors and enhance adherence, but have not demonstrated lower rates of stroke or bleeding compared to usual care, and their cost-effectiveness is unknown. We evaluated four anticoagulant strategies for patients with atrial fibrillation initiating therapy: warfarin AMS, DOAC usual care, DOAC population management tool (PMT), and DOAC AMS.

**Methods:** We developed a Markov model with monthly cycles simulating lifetime risk of ischemic stroke, major bleeding, death, disability, and costs from a US healthcare sector perspective. Costs and outcomes were discounted 3% annually. Model probabilities were derived from a prior Kaiser Permanente comparative-effectiveness analysis. Other inputs from published literature and national data. Primary outcomes were direct healthcare costs (2025 USD), quality-adjusted life years (QALYs), and incremental cost-effectiveness ratios (ICERs). Sensitivity analyses assessed parameter uncertainty.

**Results:** DOAC-based strategies yielded greater QALYs than warfarin AMS and were cost-effective at standard willingness-to-pay thresholds. Compared with warfarin AMS, DOAC usual care gained 0.4 QALYs (ICER $89,200/QALY), DOAC PMT gained 0.6 QALYs (ICER $66,700/QALY), and DOAC AMS gained 0.6 QALYs (ICER $64,500/QALY). DOAC usual care and DOAC PMT were extendedly dominated by DOAC AMS. At $120,000/QALY, DOAC AMS was preferred in 50.4% of probabilistic iterations, DOAC PMT in 36.3%, DOAC usual care in 11.0%, and warfarin AMS in 2.3%. Results were most sensitive to DOAC program effectiveness and DOAC costs.

**Conclusions:** Pharmacist-led DOAC management is cost-effective compared with warfarin AMS for AF patients. These findings support broader adoption of structured DOAC management programs to optimize anticoagulation therapy.

## INTRODUCTION

US spending on atrial fibrillation (AF) and atrial flutter has increased substantially in recent years, driven largely by rising ambulatory care and medication costs. Annual spending is estimated at $30.2 billion in 2024 USD.^1^ Over the last decade, direct oral anticoagulants (DOACs) have rapidly replaced warfarin as the predominant therapy for stroke prevention in AF and now account for nearly two-thirds of prescriptions.^2–4^ Although DOACs offer advantages over warfarin, including a wider therapeutic index and no need for routine coagulation monitoring, they still require clinical oversight. Patients prescribed DOACs may benefit from follow-up that provides education, reinforces adherence, addresses drug interactions, and supports peri-procedural management.

Pharmacist-led anticoagulation management services (AMS), often referred to as warfarin or Coumadin clinics, have long played a central role in providing high-quality anticoagulation care. In warfarin therapy, AMS collaborate closely with prescribing clinicians to perform routine coagulation monitoring, including international normalized ratio (INR) testing, adjust doses, and provide patient education. Because DOACs do not require INR monitoring or frequent titration, it is unclear whether programs designed for warfarin are necessary or beneficial for DOACs. Nevertheless, as DOACs have become first-line therapy, many health systems have expanded their anticoagulation programs to include patients using DOACs.^5^

Although DOAC management programs vary across health systems, many share features that can be broadly categorized into two care models. In this analysis, we use the term DOAC AMS to describe programs in which AMS pharmacists assume responsibility for key aspects of DOAC-related care, including initial appropriateness review, patient education, medication management, and follow-up tailored to clinical need. In contrast, we use the term DOAC population management tool (DOAC PMT) to describe approaches that rely on electronic health record–triggered reports or alerts, enabling pharmacists to intervene only when safety issues or care gaps are identified.

Kaiser Permanente (KP) provides a useful example of how health systems have implemented DOAC management programs in practice. Across its regions, KP adopted different strategies for DOAC care, including DOAC AMS, DOAC PMT, or usual care without a dedicated DOAC program. In our prior analysis of more than 44,000 KP patients with AF, this natural variation allowed us to compare outcomes across care models. In regions that implemented a DOAC management program, whether DOAC AMS or DOAC PMT, DOAC users experienced significantly fewer anticoagulation-related adverse events, defined as a composite of thromboembolic stroke, intracranial hemorrhage, other major bleeding, or death, than warfarin users managed through a traditional AMS (hazard ratio [HR], 95 percent confidence interval [CI]: DOAC AMS vs warfarin AMS, 0.84 [0.72 to 0.99]; DOAC PMT vs warfarin AMS, 0.85 [0.79 to 0.90]). In contrast, this difference was not significant in the region that did not implement a DOAC program (HR, 95 percent CI: DOAC usual care vs warfarin AMS, 0.91 [0.79 to 1.05]). Among patients receiving DOACs, outcomes were similar between regions using DOAC AMS and those using DOAC PMT.

Health systems frequently consider adopting DOAC AMS or PMT-based care models, yet data are limited on the costs of these services, their potential value, and the magnitude of clinical benefit required for them to be cost-effective. Economic modeling can clarify the conditions under which these programs would represent good value, identify the degree of outcome improvement needed to justify investment, and inform decisions about the design and implementation of DOAC management strategies. In this analysis, we used a computer simulation model to estimate the lifetime health outcomes, costs, and cost-effectiveness of DOAC management strategies, including DOAC AMS, DOAC PMT, and DOAC usual care, compared with warfarin AMS from a US healthcare sector perspective.

## METHODS

### Setting

The comparative effectiveness study underlying this economic analysis has been described previously. Briefly, it was a retrospective cohort analysis conducted within KP, an integrated health care system consisting of eight regions across the United States. Our analysis included the Northwest, Southern California, and Colorado regions, which collectively served more than 5.8 million members as of March 2022. Data were obtained from the Virtual Data Warehouse, a standardized data infrastructure that harmonizes information across regions and includes demographics, diagnoses, procedures, pharmacy records, vital signs, laboratory measurements, and health care utilization.

All KP regions used pharmacist-led AMS for warfarin management. In contrast, DOAC management was not standardized at the national level and varied by region. The three regions included in this analysis were selected because each had adopted a different region-wide strategy for DOAC care, representing DOAC usual care, DOAC AMS, or DOAC PMT. This natural variation provided an opportunity to compare distinct anticoagulation management approaches as they existed in routine clinical practice.

### Anticoagulation Care Strategies

Although DOACs are now guideline-preferred for most patients with AF, warfarin remains a relevant treatment option in specific clinical situations and in settings where medication cost or access is a concern. AMS is the recommended approach for managing warfarin therapy and served as the common comparator in the underlying comparative effectiveness study from which treatment effect estimates were derived. For these reasons, warfarin AMS was used as the reference strategy in our simulation model. We compared three DOAC management models with this reference strategy: DOAC usual care, DOAC AMS, and DOAC PMT. These models reflect real-world approaches that have been implemented across large US health systems, including KP, and represent the types of programmatic decisions health systems face when designing DOAC care.

Warfarin AMS was a pharmacist-led care model in which AMS pharmacists managed all aspects of warfarin therapy under collaborative practice agreements. Core responsibilities included patient education, verification of indication and dose appropriateness, ordering and interpretation of laboratory tests such as renal function and INR, dose adjustment based on laboratory and clinical findings, and longitudinal follow-up. Pharmacists typically met with patients at regular intervals, reviewed anticoagulation control and adherence, and coordinated care with prescribing clinicians.

DOAC usual care was a clinician-led model in which the DOAC prescriber managed all aspects of DOAC therapy. No structured pharmacist involvement or systematic DOAC-specific monitoring protocol was included in this approach.

DOAC AMS extended the pharmacist-led AMS model used for warfarin to DOAC therapy. AMS pharmacists managed all DOAC users under collaborative practice agreements and provided patient education, reviewed indication and dose appropriateness, assessed potential drug interactions, ordered relevant laboratory tests such as renal function, and provided longitudinal follow-up.

DOAC PMT supplemented usual clinician-directed care with a pharmacist-managed, EHR-based population management tool. The PMT automatically flagged patients with potential DOAC-related concerns such as inappropriate dose or indication, drug–drug interactions, medication nonadherence, overdue or abnormal laboratory results, or upcoming invasive procedures. Pharmacists reviewed these flagged cases and intervened when appropriate by contacting the prescribing clinician or the patient. Unlike DOAC AMS, DOAC PMT did not involve routine or proactive pharmacist follow-up for all DOAC users; pharmacist involvement occurred only in response to PMT-generated alerts.

### Model Overview

We developed a Markov health state–transition model to simulate the lifetime clinical and economic consequences of anticoagulation therapy for individuals with AF (**Figure 1**, **Supplemental Figure 1**, and **Supplemental Table 1**). The simulated cohort matched the mean characteristics of patients with AF initiating anticoagulation therapy in the three KP regions previously described.^6^ The baseline mean age was 73 years and 43.9 percent were female. All individuals entered the simulation at the time they initiated oral anticoagulation therapy and were assumed to have no chronic disability at baseline.

**Figure 1.**
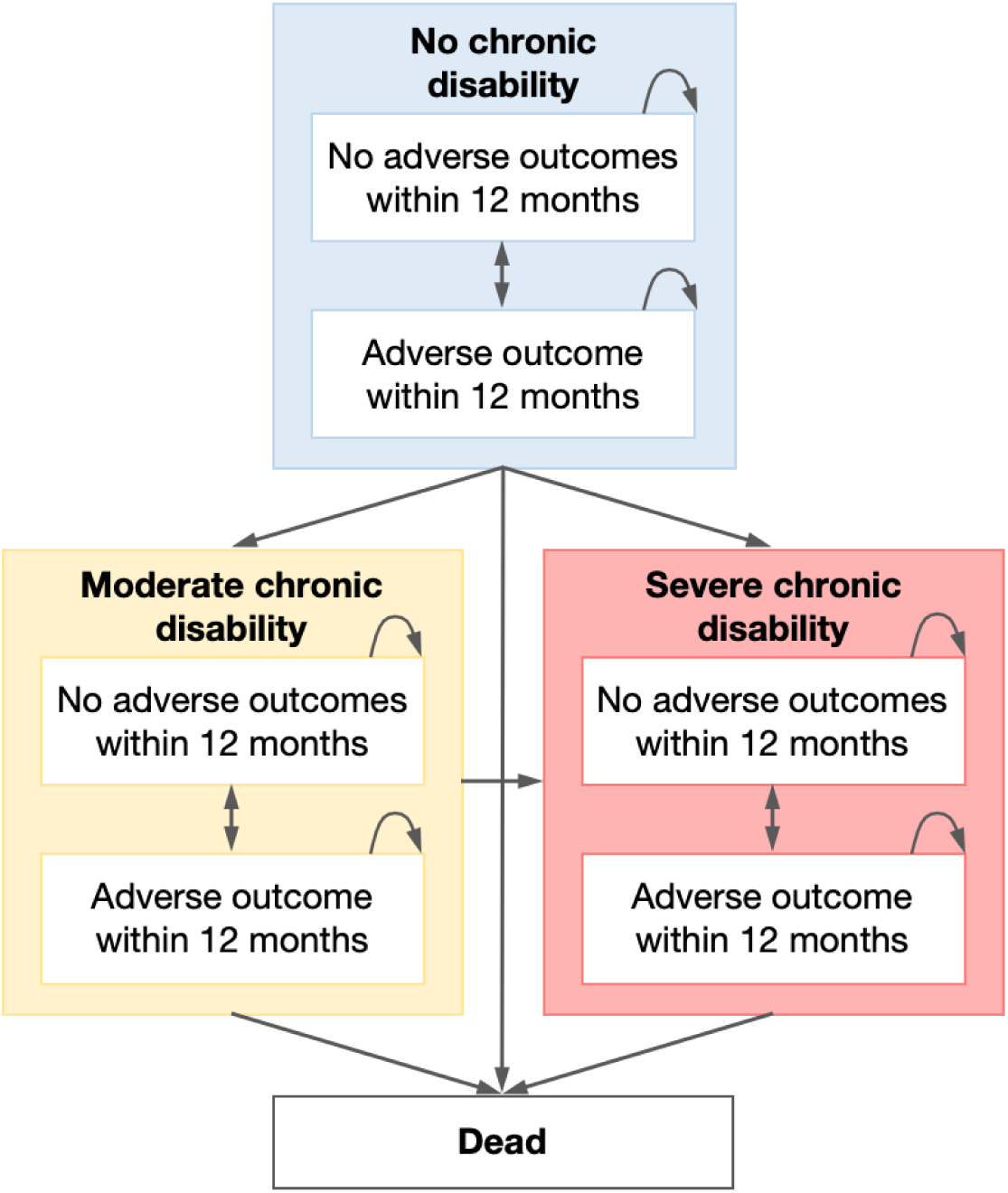
Markov model schematic. Notes: Arrows represent allowed transitions between health states during each model cycle. Transition probabilities determine movement between states.

The model used monthly cycles. In each cycle, individuals could remain event-free or experience one of the following adverse outcomes: (1) fatal or nonfatal ischemic stroke, (2) fatal or nonfatal major bleed, categorized as intracranial hemorrhage (ICH) or extracranial hemorrhage (ECH), or (3) death not attributable to stroke or major bleeding. Following an ischemic stroke or ICH, individuals could transition to moderate or severe chronic disability. ECH events were assumed not to lead to long-term disability. Health-related quality of life and health care costs were updated each month based on adverse outcomes experienced, time since the most recent event, and current disability level.

### Model Parameters

#### Probability of events

The probability of experiencing an adverse outcome under warfarin AMS was derived from the comparative effectiveness study conducted within KP (**Table 1**, **Supplemental Methods**, and **Supplemental Table 2**).^6^ We used hazard ratios from that study to estimate the relative risk of adverse outcomes associated with each DOAC management strategy compared with warfarin AMS: DOAC usual care, 0.91 (95 % CI, 0.79 to 1.05); DOAC AMS, 0.84 (95% CI, 0.72 to 0.99); DOAC PMT, 0.85 (95% CI, 0.79 to 0.90).

**Table 1.**
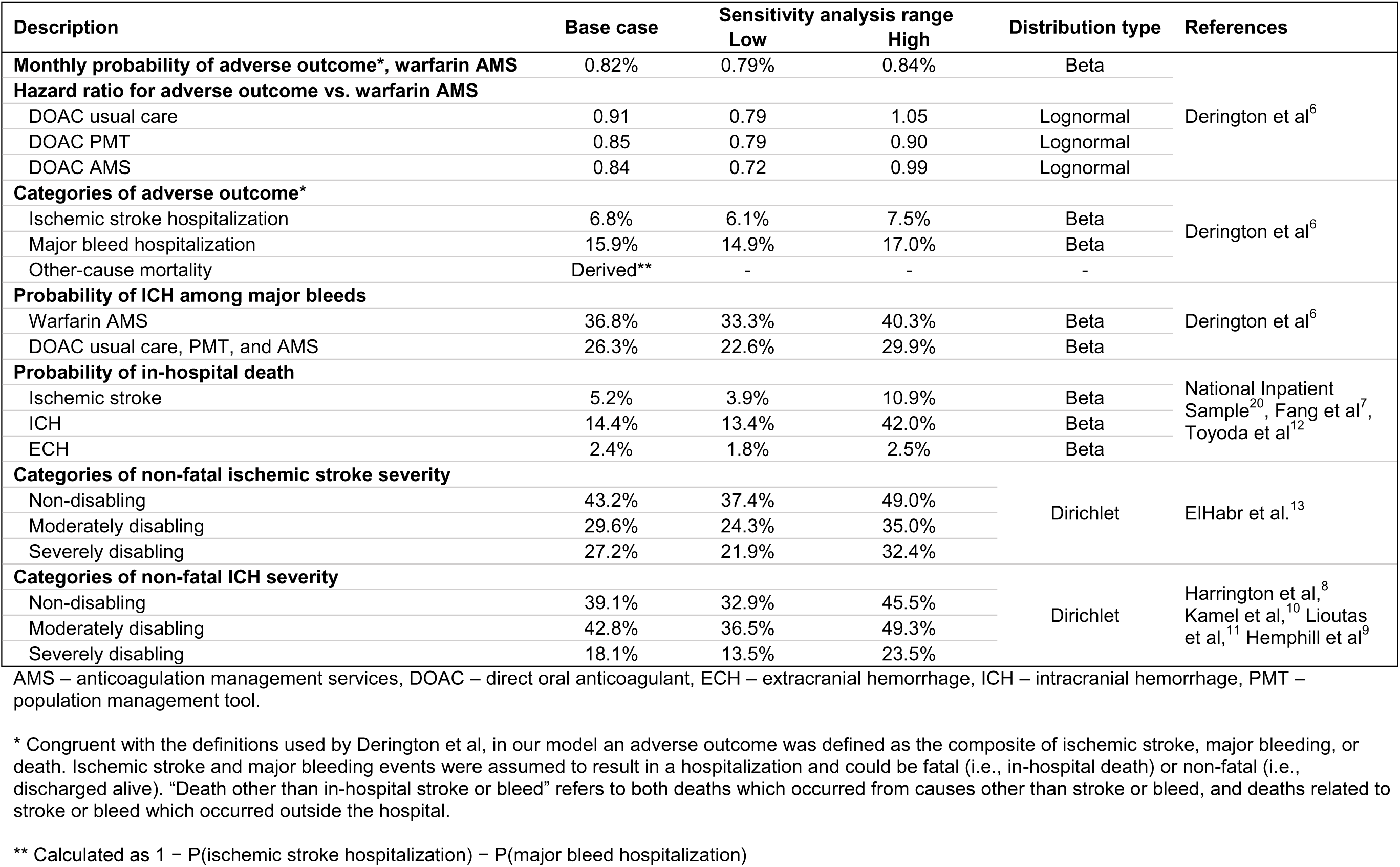
Clinical model inputs and assumed distributions.

When an adverse outcome occurred, event type (ischemic stroke, ICH, ECH, or other death) was assigned according to the distribution observed in the comparative effectiveness study. Only one adverse event could occur within a monthly cycle. Published estimates informed the probability and severity of chronic disability following ischemic stroke or ICH; ECH was assumed not to result in long-term disability.^7–13^

The median follow-up in the comparative-effectiveness analysis was 2 years, thus we assumed the probability of death was the same for the first 2 years of the simulation.^6^ To extrapolate beyond this in subsequent years, we accounted for an increased risk of death associated with aging using the 2019 Centers for Disease Control and Prevention lifetables, matching the last year of eligibility in the comparative-effectiveness study (**Supplemental Figure 2**).^14^ In the primary analysis, we assumed the effects of the DOAC management strategies were sustained for the remaining lifetime.

#### Costs

Cost inputs were inflated to 2025 US Dollars ($) using the healthcare component of the Personal Consumption Expenditures Index from the US Bureau of Economic Analysis when necessary (**Table 2**). Medication costs were from the Veterans Affairs Federal Supply Schedule.^15,16^ We assumed an average warfarin dose of 5 mg daily and calculated the monthly cost as the simple average of 5 mg tablets. We calculated the average monthly price of DOACs as the simple average of apixaban, dabigatran, edoxaban, and rivaroxaban, assuming perfect adherence (i.e., the medication was purchased every month). For warfarin, we assumed two INR tests per month, with costs derived from the Centers for Medicare and Medicaid Services.^17^

**Table 2.**
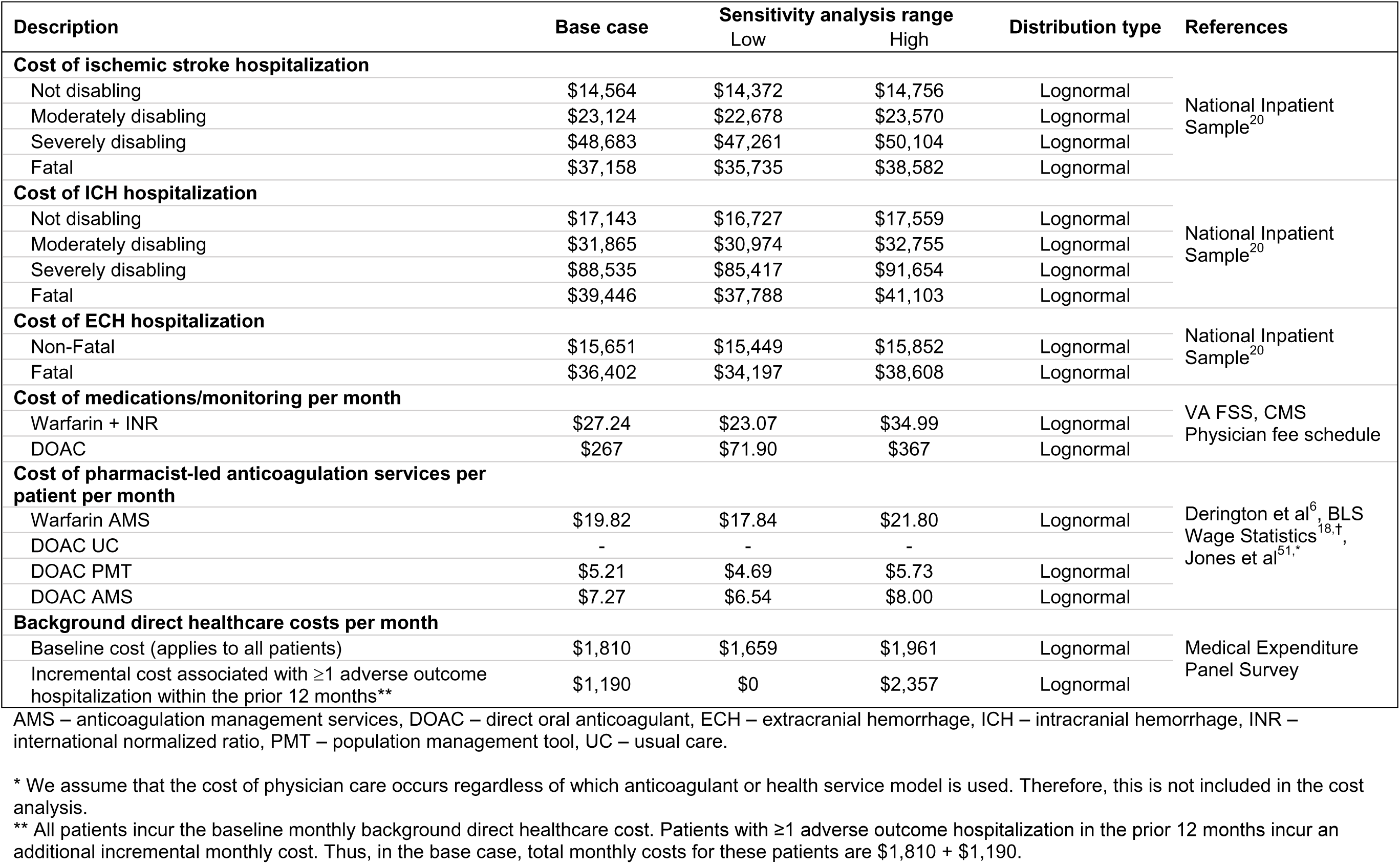
Model inputs for costs (2025 USD) and assumed distributions.

Warfarin AMS, DOAC PMT, and DOAC AMS were assumed to be provided by pharmacists and pharmacy technicians. Labor costs were derived from goal staffing ratios (**Supplemental Methods**) and national annual wage estimates.^18^ Adverse outcomes were assumed to result in a hospitalization, and total hospitalization costs were derived from the 2018 National Inpatient Sample and a professional fee ratio from the published literature (**Supplemental Methods**).^19,20^ Background healthcare costs for adults with AF were estimated using total expenditures from the 2016-2023 Medical Expenditure Panel Survey, with costs of anticoagulant medications and adverse outcome hospitalizations subtracted. Background healthcare costs were assumed to be higher during the first year after an adverse outcome (**Supplemental Methods**).^21^ Time since the most recent adverse outcome was used only to classify individuals into background cost categories (<12 months vs. ≥12 months). The corresponding cost estimates were then applied monthly, consistent with all other model inputs.

#### Health-related Quality of Life

We used a quality-of-life weight of 0.81 for individuals with AF who are otherwise healthy (i.e., no chronic disability).^22^ We applied quality-of-life decrements associated with increasing age, medication use, and chronic moderate or severe disability that resulted from an ischemic stroke or intracranial hemorrhage (**Table 3**).^23–26^ We also applied a one-time quality-of-life decrement associated with adverse outcomes at the time the event occurred.

**Table 3.**
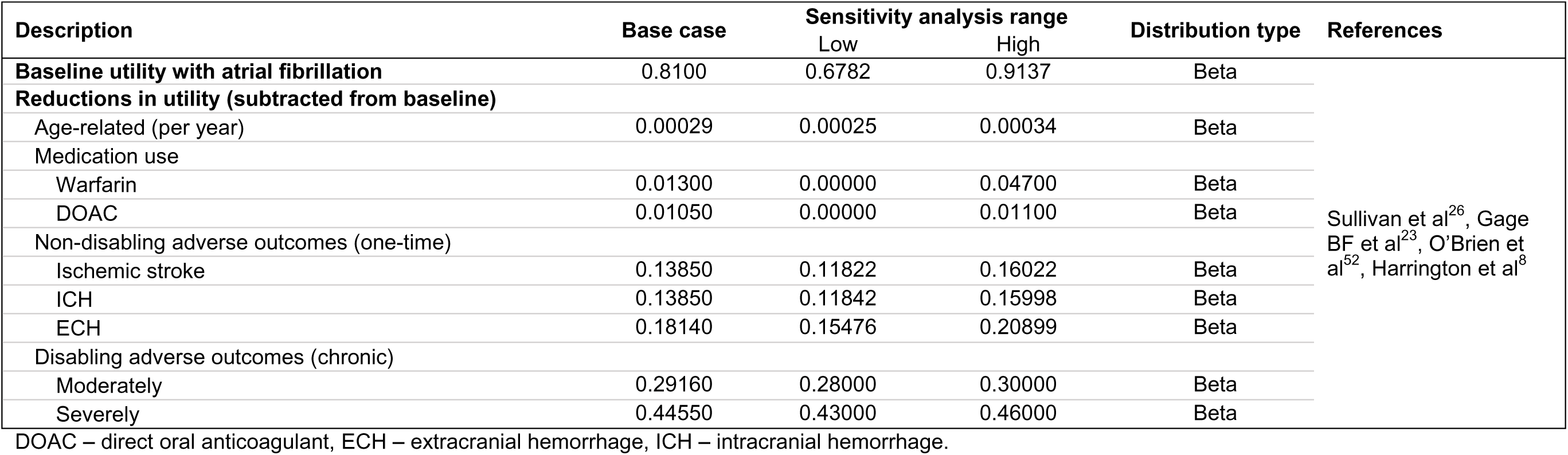
Model inputs for quality of life and assumed distributions.

### Statistical Analysis

Our analyses and reporting adhered to recommendations from the Second Panel on Cost-effectiveness in Health and Medicine (**Supplemental Table 3**).^27^

The primary outcomes were quality-adjusted life years (QALYs), which adjust survival for health-related quality of life, and direct healthcare costs (2025 US Dollars). They were then used to calculate cost-effectiveness, measured by the incremental cost-effectiveness ratio (ICER) which was expressed as the cost per QALY gained compared with warfarin AMS. We used the value framework from the American College of Cardiology and American Heart Association to classify ICERs as cost-effective when <$120,000 per QALY gained.^28^ Secondary outcomes included survival, medication costs, anticoagulation services (e.g., pharmacist time, laboratory monitoring), adverse outcomes, and “background” healthcare utilization (i.e., direct healthcare costs incurred not related to anticoagulation medications or adverse outcome hospitalizations).

All analyses were performed from a healthcare sector perspective and future costs and QALYs were discounted at an annual rate of 3% (varied from 1% to 8% in sensitivity analysis) (**Supplemental Table 4**).^27^ The mean and 95% uncertainty interval (UI), i.e., 2.5^th^ to 97.5^th^ percentile, were calculated for key outcomes by running 1000 iterations of the model in which model parameters were randomly sampled from statistical distributions. Monthly probabilities and event-specific probabilities were modeled using beta distributions. Hazard ratios were modeled using lognormal distributions. Severity categories for non-fatal stroke and intracranial hemorrhage were modeled using Dirichlet distributions to preserve the correlation structure across mutually exclusive outcome categories. Cost parameters were modeled using lognormal distributions to reflect right-skewed cost data. The model was developed and all simulation analyses were performed using TreeAge Pro 2025 (TreeAge Software Inc, Williamstown, MA). Additional analyses were performed using Stata (version 15.1) and R (version 4.5.1; Vienna, Austria). A strategy was considered preferred if it resulted in the greatest QALY gains and had an ICER less than the willingness-to-pay threshold.

### Sensitivity and scenario analyses

One-way and two-way sensitivity analyses were conducted to explore the effect of parameter uncertainty on cost-effectiveness estimates. One-way sensitivity analysis included DOAC medication costs and incorporated the negotiated prices for rivaroxaban and apixaban established by the Centers for Medicare and Medicaid Services under the Inflation Reduction Act of 2022, referred to as the “Maximum Fair Price.” In the comparative effectiveness analysis, about 50% of individuals discontinued DOACs in the first year. We therefore included in the one-way sensitivity analysis a variable that reduced medication costs and medication use associated quality-of-life decrement by up to 50%, assuming that the HRs were intention-to-treat effects. Additionally, two scenario analyses were conducted to assess the long-term effectiveness of each DOAC treatment strategy. In the first scenario, the HR for each DOAC treatment strategy decreased linearly over 10 years after the first 2 years until there was no difference with warfarin AMS (i.e., HR = 1 at 12 years). In the second scenario, the duration of the effectiveness (i.e., time until the HR = 1) was varied.

## RESULTS

In the base-case analysis, DOAC-based strategies were associated with longer survival and greater QALYs than warfarin AMS, at higher overall healthcare costs; DOAC AMS was the most cost-effective strategy at a willingness-to-pay threshold of $120,000 per QALY.

### Model Validation and Base-Case Analysis

At 2 years, the model reproduced the adverse outcome event rates and HRs from the comparative effectiveness analysis (**Supplemental Table 5**).^6^ The projected proportion of the population surviving at 13 years was similar to the proportion of participants with AF from the Framingham Heart Study (**Supplemental Figure 2**).^29^

The projected mean undiscounted survival was 8.9 years (95% UI: 8.8 - 9.1) with warfarin AMS, 9.6 years (95% UI: 8.7 - 10.5) with DOAC usual care, 10.0 years (95% UI: 9.5 - 10.5) with DOAC PMT, and 10.1 years (95% UI: 9.2 - 11.2) with DOAC AMS (**Table 4**). Compared with warfarin AMS, the number of adverse outcomes per 100,000 individuals treated over the remaining lifetime was projected to decrease by 274 (95% UI: −598 to 1493) with DOAC usual care, 747 (95% UI: 40 to 1450) with DOAC PMT, and 927 (95% UI: −271 to 2667) with DOAC AMS. Warfarin AMS was projected to yield a mean of 5.7 QALYs (95% UI: 4.8 to 6.5), which was projected to increase by 0.4 QALYs (95% UI: −0.1 to 0.9) with DOAC usual care, 0.6 QALYs (95% UI: 0.3 to 0.9) with DOAC PMT, and 0.6 QALYs (95% UI: 0.1 to 1.3) with DOAC AMS.

**Table 4.** Cost-effectiveness outcomes.

| Outcomes | Warfarin AMS | DOAC Usual Care | DOAC PMT | DOAC AMS |
| --- | --- | --- | --- | --- |
| **Effectiveness** |  |  |  |  |
| Life years (undiscounted) | 8.9 (8.8 - 9.1) | 9.6 (8.7 - 10.5) | 10.0 (9.5 - 10.5) | 10.1 (9.2 - 11.2) |
| Incremental life years (undiscounted) | Reference | 0.6 (-0.2 - 1.6) | 1.1 (0.7 - 1.5) | 1.2 (0.2 - 2.3) |
| Life years (discounted) | 7.4 (7.3 - 7.5) | 7.8 (7.2 - 8.5) | 8.1 (7.8 - 8.5) | 8.2 (7.5 - 9.0) |
| Incremental life years (discounted) | Reference | 0.4 (-0.2 - 1.1) | 0.8 (0.5 - 1.1) | 0.8 (0.2 - 1.6) |
| QALYs | 5.7 (4.8 - 6.5) | 6.1 (5.0 - 7.1) | 6.3 (5.3 - 7.3) | 6.4 (5.2 - 7.5) |
| Incremental QALYs | Reference | 0.4 (-0.1 - 0.9) | 0.6 (0.3 - 0.9) | 0.6 (0.1 - 1.3) |
| **Direct Healthcare Costs (2025 USD)** |  |  |  |  |
| Total | \$191,762 (\$163,305 - \$293,169) | \$223,887 (\$183,766 - \$319,876) | \$232,009 (\$193,107 - \$324,320) | \$233,653 (\$191,070 - \$333,080) |
| Increase in total | Reference | \$32,125 (\$9,387 - \$64,225) | \$40,247 (\$21,401 - \$72,406) | \$41,891 (\$18,787 - \$74,557) |
| Anticoagulation services | \$1,802 (\$1,610 - \$1,993) | \$0 (\$0 - \$0) | \$518 (\$465 - \$579) | \$732 (\$637 - \$836) |
| Increase in anticoagulation services | Reference | -\$1,802 (-\$1,993 - -\$1,610) | -\$1,283 (-\$1,473 - -\$1,087) | -\$1,070 (-\$1,273 - -\$863) |
| Medication | \$2,479 (\$2,013 - \$3,064) | \$27,731 (\$11,461 - \$56,758) | \$28,807 (\$11,791 - \$58,773) | \$29,020 (\$11,816 - \$59,103) |
| Increase in medication | Reference | \$25,251 (\$8,848 - \$54,126) | \$26,328 (\$9,244 - \$56,555) | \$26,541 (\$9,255 - \$57,063) |
| Adverse outcomes | \$5,511 (\$5,082 - \$5,968) | \$5,093 (\$4,678 - \$5,517) | \$5,019 (\$4,641 - \$5,407) | \$5,003 (\$4,590 - \$5,431) |
| Increase in adverse outcomes | Reference | -\$419 (-\$652 - -\$195) | -\$492 (-\$681 - -\$291) | -\$509 (-\$751 - -\$274) |
| Background | \$181,969 (\$153,499 - \$282,406) | \$191,063 (\$157,883 - \$288,964) | \$197,664 (\$166,480 - \$295,191) | \$198,898 (\$164,430 - \$304,025) |
| Increase in background | Reference | \$9,094 (-\$3,570 - \$22,998) | \$15,695 (\$9,479 - \$22,395) | \$16,928 (\$3,307 - \$33,436) |
| **Cost-effectiveness** |  |  |  |  |
| ICER vs. warfarin AMS (\$ per QALY gained) | Reference | \$89,100* | \$66,700* | \$64,500 |
| Preferred strategy at \$120,000/QALY | 2.3% | 11.0% | 36.3% | 50.4% |

AMS – anticoagulation services, DOAC – direct oral anticoagulant, ICER – incremental cost-effectiveness ratio, PMT – population management tool, QALYs – quality-adjusted life years, USD – US dollars.
\*Strategies were extendedly dominated by DOAC AMS and represent inefficient care (i.e., gained fewer QALYs than DOAC AMS at a higher cost per QALY gained); therefore, DOAC AMS would be preferred.
Notes: The table shows the projected effectiveness, cost, and cost-effectiveness outcomes when running 1000 iterations of the model and randomly sampling all model inputs from prespecified statistical distributions for each iteration. The values are presented as the mean and 95% uncertainty interval (2.5<sup>th</sup> to 97.5<sup>th</sup>).
percentile) for each outcome. The probability that each management strategy was preferred is the proportion of iterations in which it resulted in the greatest QALY gains and had an ICER less than the cost-effectiveness threshold ( $< \$120,000$ per QALY).

Estimated mean direct healthcare costs were $191,762 (95% UI: $163,305 - $293,169) with warfarin AMS, with estimated increases of $32,125 (95% UI: $9,387 - $64,225) with DOAC usual care, $40,247 (95% UI: $21,401 - $72,406) with DOAC PMT, and $41,891 (95% UI: $18,787 - $74,557) with DOAC AMS. Estimated costs saved by avoiding adverse outcomes with DOAC management strategies were offset by increased medication costs and costs due to increased survival. Compared with warfarin AMS, DOAC AMS was cost-effective (i.e., ICER <$120,000 per QALY gained) with an ICER of $64,500 per QALY gained. DOAC usual care ($89,100 per QALY gained vs. warfarin AMS) and DOAC PMT ($66,700 per QALY gained vs. warfarin AMS) were extendedly dominated by DOAC AMS (i.e., representing less efficient care, gaining fewer QALYs than DOAC AMS at a higher cost per QALY gained). At lower willingness-to-pay thresholds (<$50,000 per QALY), warfarin AMS had the highest probability of being preferred; however, this probability declined rapidly as the threshold increased (**Figure 2**). At a contemporary cost-effectiveness threshold of $120,000 per QALY, DOAC AMS had the highest probability of being preferred (50.4%), followed by DOAC PMT (36.3%), DOAC usual care (11.0%), and warfarin AMS (2.3%).

**Figure 2.**
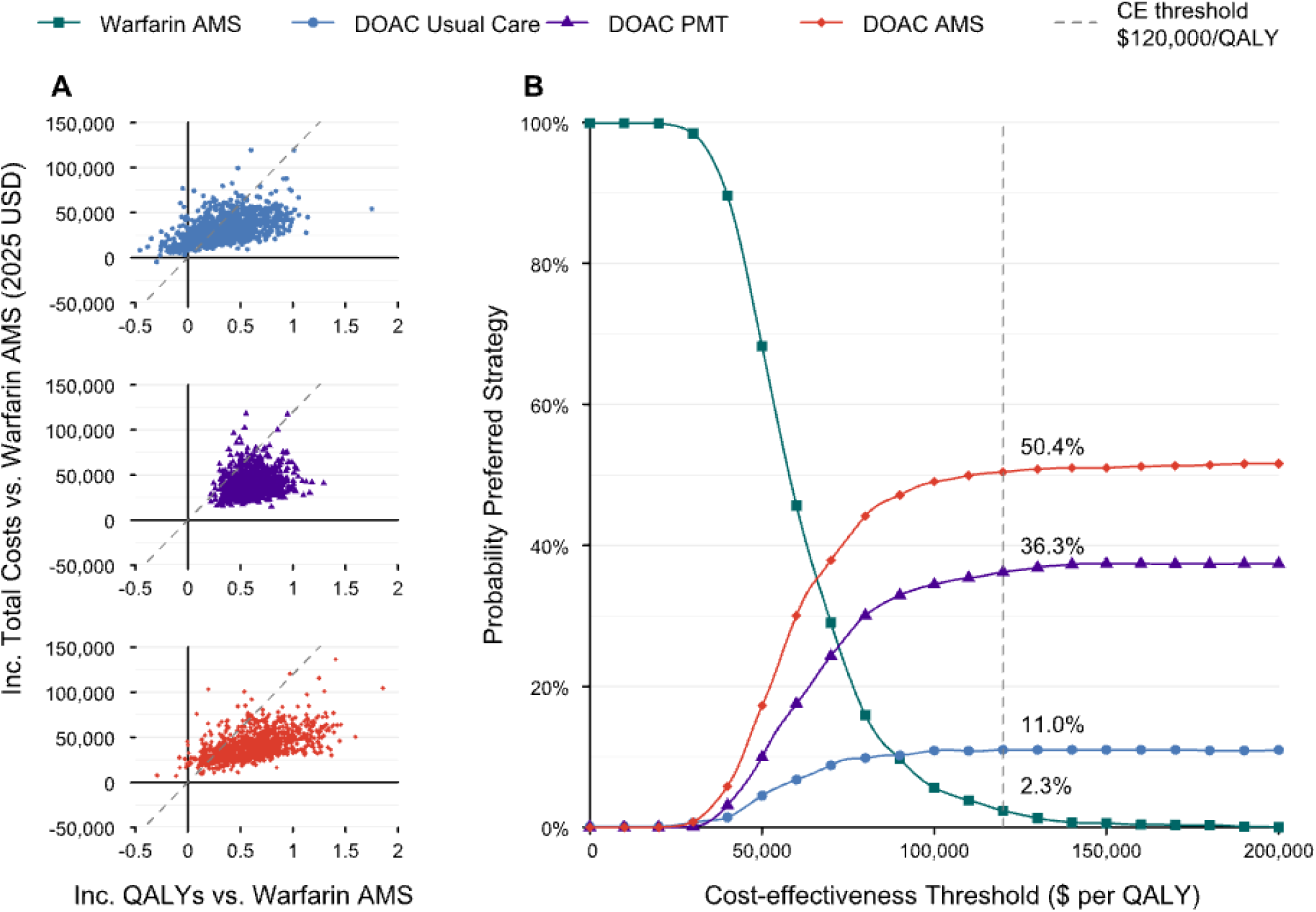
Incremental cost-effectiveness scatterplots and cost-effectiveness acceptability curve. AMS – anticoagulation services, CE – cost-effectiveness, DOAC – direct oral anticoagulant, PMT – population management tool, QALYs – quality-adjusted life years, USD – US dollars. Notes: The figure shows the incremental total direct healthcare costs and incremental QALYs of DOAC management strategies compared with warfarin AMS in **Panel A** and the probability that each management strategy was preferred (i.e., resulted in the greatest QALY gains and had an incremental cost-effectiveness ratio less than the cost-effectiveness threshold) across a range of cost-effectiveness thresholds in **Panel B**. The grey dashed lines represent the likely cost-effectiveness threshold for payers in the US of $120,000 per QALY. The results are from running 1000 iterations of the model and randomly sampling all model inputs from prespecified statistical distributions for each iteration. Each point in Panel A represents the results from one iteration. The points below the line represent iterations in which the strategy was considered cost-effective vs. warfarin AMS (i.e., the incremental cost-effectiveness ratio was <$120,000 per QALY). The probabilities in Panel B are the proportion of iterations in which the strategy was preferred at that cost-effectiveness threshold.

### Sensitivity and Scenario Analyses

The ICERs were most sensitive to the HR for adverse outcomes vs. warfarin AMS and the monthly cost of DOACs (**Figure 3**). When the HR for adverse outcomes vs. warfarin AMS was less than 0.95, the DOAC management strategies were projected to be cost-effective vs. warfarin AMS (i.e., ICERs <$120,000 per QALY gained) (**Supplemental Figure 3**). When the monthly cost of DOACs was >$100, DOAC usual care and DOAC PMT were extendedly dominated by DOAC AMS, and DOAC AMS remained cost-effective vs. warfarin AMS even when the monthly cost of DOACs was $500 (**Supplemental Figure 4**).

**Figure 3.**
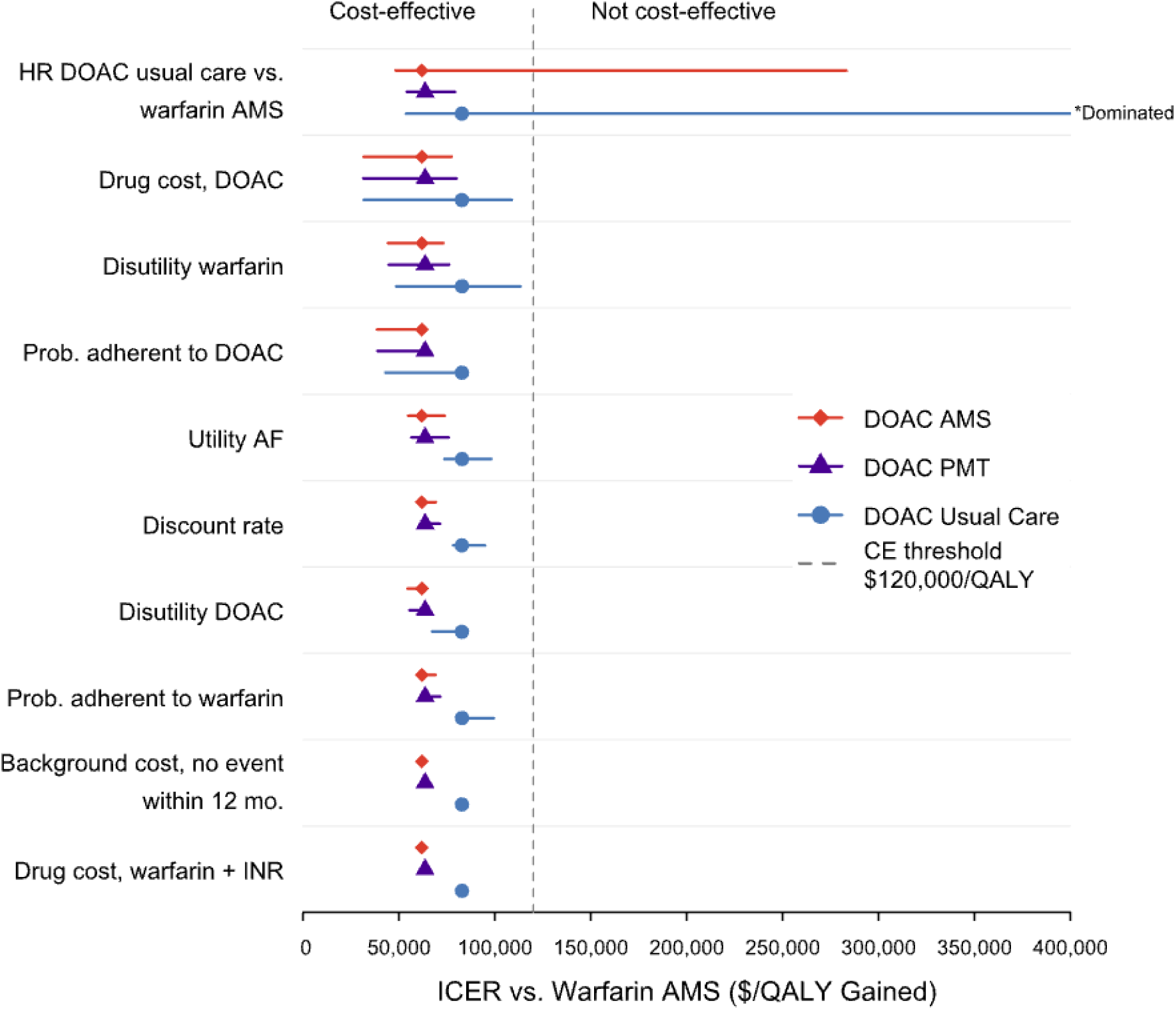
One-way sensitivity analysis. AF – atrial fibrillation, AMS – anticoagulation management services, CE – cost-effectiveness, DOAC – direct oral anticoagulant, HR – hazard ratio, ICER – incremental cost-effectiveness ratio, INR – international normalized ratio, mo. – months, PMT – population management tool, Prob. – probability, QALYs – quality-adjusted life years, USD – US dollars. Notes: The figure shows how the ICER vs. warfarin AMS changed for each DOAC management strategy as model inputs were independently varied (all other inputs held at base-case value) across a plausible range. Only the 10 most influential model inputs are shown. The base-case ICER for each strategy is represented by the points and the lines indicate the change in the ICER. The grey dashed line represents the likely cost-effectiveness threshold of US payers, $120,000 per QALY gained – values to the left of the line indicate the strategy was cost-effective vs. warfarin AMS.

The projected number of adverse outcomes and ICERs were sensitive to the time horizon (**Supplemental Figure 5**). Due to a longer projected life expectancy, more adverse outcomes were projected to occur with DOAC strategies later in life. Compared with warfarin AMS up to age 85 years, the number of adverse outcomes per 100,000 individuals was projected to decrease by 4189 (95% UI: −2037 to 11,070) with DOAC usual care, 7439 (95% UI: 4604 to 10,672) with DOAC PMT, and 8039 (95% UI: 1276 to 15,814) with DOAC AMS. The two-way sensitivity analyses projected that HR of adverse outcomes vs. warfarin AMS had a larger effect on ICERs than the cost of DOAC management services (**Supplemental Figures 6** and **7**).

In a scenario analysis that incorporated 50% discontinuation of DOACs in the first year, with corresponding lower drug costs, the ICER of DOAC AMS vs. warfarin AMS decreased to $38,514 (**Supplemental Table 6**). DOAC AMS was cost-effective and remained the preferred strategy in the scenario analyses that varied the duration of benefit with DOAC management over time (**Supplemental Table 7** and **Supplemental Figure 8**).

## DISCUSSION

In this simulation analysis of patients with AF, we projected the lifetime healthcare costs and QALYs of DOAC management strategies compared with warfarin AMS. We estimated that DOAC AMS and DOAC PMT would both gain 0.6 QALYs compared with warfarin AMS, at a cost of $56,100 and $57,100 per QALY gained, respectively. Both DOAC AMS and DOAC PMT were estimated to be intermediate value strategies using the American College of Cardiology and American Heart Association value framework. The cost-effectiveness of DOAC management strategies were sensitive to the risk of adverse outcomes and medication costs.

To our knowledge, this is the first study to evaluate the cost-effectiveness of various approaches to providing longitudinal management of DOAC therapy in patients with AF. Prior to the DOAC era, Sullivan et al. estimated that warfarin AMS cost less and was more effective than usual care (i.e., physician management of warfarin therapy).^26^ When DOACs became commercially available in 2010, many assumed that structured management would not be necessary because DOACs do not require routine laboratory monitoring like warfarin. Under this assumption, DOACs were estimated to be cost-effective vs. warfarin.^30^ However, real-world experience has highlighted persistent challenges in DOAC use, including dosing errors, drug-drug interactions, and poor patient adherence, all of which result in adverse outcomes.^31–38^ Indeed, even with DOACs supplanting warfarin as the most common oral anticoagulant, emergency visits for oral anticoagulant-associated harm remain virtually unchanged.^39–41^ These findings suggest that the absence of routine laboratory monitoring does not eliminate the need for structured clinical oversight. Health systems must determine whether investment in DOAC-focused programs or technologies are financially justified.

In our analysis, all DOAC strategies were cost-effective compared to warfarin AMS. However, DOAC usual care and DOAC PMT gained fewer QALYs at a higher cost than DOAC AMS, which is referred to as extended dominance. Extendedly dominated strategies (DOAC usual care and DOAC PMT in this analysis) are often ruled out of consideration given a preference for more effective interventions with lower ICERs. However, from a practical standpoint, DOAC PMT and DOAC AMS yielded similar gains in QALYs and healthcare costs making it difficult to use the results of this analysis alone for choosing one strategy over the other. Any DOAC management strategy needs to be integrated into existing healthcare frameworks and clinical workflows. The preferred approach may depend on local context, infrastructure, and personnel resources needed for implementation, which could be informed by future qualitative and cost-effectiveness analyses. Although we did not calculate ICERs for DOAC AMS vs DOAC PMT or DOAC usual care, as is standard practice with dominated or extendedly dominated strategies, it should be noted that, by definition, the ICER would be lower than the ICER of DOAC AMS vs warfarin AMS. Therefore, even if warfarin was removed from consideration, AMS would remain a cost-effective strategy to manage DOACs.

Although our primary analysis projected outcomes over a lifetime horizon, health systems may evaluate program investments over shorter time frames that align with patient retention within a given insurance plan. Turnover in US commercial health insurance is common, with roughly one in five enrollees leaving their plan each year and average enrollment durations of approximately four years.^42^ This consideration is less relevant for Medicare beneficiaries, who make up a majority of DOAC users; while switching between Medicare Advantage plans occurs, beneficiaries remain within the broader Medicare program, which ultimately bears the cost of major adverse events regardless of the specific plan.^43^ In time-horizon sensitivity analyses, the ICER for DOAC AMS and DOAC PMT fell below $120,000 per QALY gained at approximately 5 years, suggesting that structured DOAC management may reach commonly cited willingness-to-pay thresholds within a timeframe relevant to health system decision-making. Beyond this point, cost-effectiveness continues to improve as reductions in stroke and major bleeding accumulate over time.

Anticoagulation programs and services, both for DOACs and warfarin, will continue to evolve as new research and technology emerge. Wearable devices, mobile health apps, telemedicine services, pharmacogenomics, artificial intelligence and others have the potential to change the landscape of oral anticoagulant management. Guo et al, showed that using a mobile health technology with AF screening and an integrated management strategy—anticoagulation, AF symptom management, and cardiovascular risk management—can reduce adverse clinical outcomes.^44^ There are on-going efforts within the Veterans Health Administration to change the current DOAC care model paradigm, by leveraging electronic health records and a population health approach to optimize the safety and efficacy of DOACs as efficiently as possible.^45–47^ As new innovations are tested and implemented, it will be necessary to ensure that these approaches provide both clinical and economic benefits compared to established management models.

This study has several limitations to consider. Although widely used in cost-effectiveness analyses, simulation models such as Markov models necessarily simplify complex situations. No model can capture all nuances of patient management. For example, our primary analysis did not consider medication discontinuation, and while medication discontinuation was considered in sensitivity analysis, we did not account for re-initiation of anticoagulation therapy in subsequent years. With a few exceptions, DOACs are recommended as the first-line therapy over warfarin in patients with AF.^48^ Nonetheless, we chose to include warfarin as a comparator because it is a viable anticoagulation option, particularly in frail, elderly, valvular AF, mechanical heart valves, and when cost is a concern.^49^ We only evaluated warfarin therapy as managed by AMS, as opposed to warfarin managed in other settings, because prior research has shown that AMS yield more time spent within therapeutic range and have improved clinical outcomes at lower cost compared to usual care.^26,50^ Our model probabilities are based on real-world experience in the Kaiser Permanente health system, which is the largest analysis on DOAC management to date. Nonetheless, no randomized trial has been conducted to definitively show the impact of DOAC management programs on clinical outcomes. As an integrated delivery system with established population management infrastructure, Kaiser Permanente may implement anticoagulation programs more efficiently than less integrated or resource-limited healthcare settings. Event rates, implementation costs, and workflow integration may differ in other systems, which could influence cost-effectiveness estimates. Additionally, health systems may design management programs that differ from those evaluated in this analysis, potentially leading to different clinical and economic outcomes. Our model assumed that management programs primarily influence the probability of adverse events but not their severity if events occur. However, these programs could plausibly facilitate earlier detection of complications, more rapid clinical intervention, or mitigation of drug interactions, which may reduce the severity or downstream consequences of stroke or bleeding events. If so, our analysis may underestimate the potential clinical and economic benefits of structured DOAC management. Our base-case analysis assumed that the benefits of DOAC management were sustained over a patient’s lifetime, which may overestimate long-term effects. However, sensitivity analyses demonstrated that cost-effectiveness thresholds were reached within approximately five years, and DOAC AMS and DOAC PMT remained below willingness-to-pay thresholds even when the duration of treatment effect was limited to 2 years, consistent with the median follow-up in our real-world cohort analysis.^6^ Further research is needed to confirm the long-term durability of these programs in real-world settings.

## CONCLUSION

In this simulation analysis of patients with AF, pharmacist-led DOAC management services were estimated to be a cost-effective strategy to reduce adverse outcomes and increase survival compared with warfarin AMS. As new services and technologies evolve, including the introduction of new oral anticoagulants into the market, further research will be needed to assess their impact on anticoagulation management quality and financial outcomes.

## Data Availability

No individual-level patient data were used in this analysis. The analysis was conducted using a decision-analytic Markov model populated with parameters derived from published literature and publicly available sources. All model inputs are reported in the manuscript and/or supplementary materials.

## Non-standard abbreviations and acronyms

AF: Atrial fibrillation
AMS: Anticoagulation management service
CI: Confidence interval
DOAC: Direct oral anticoagulants
ECH: Extracranial hemorrhage
ICER: Incremental cost-effectiveness ratio
ICH: Intracranial hemorrhage
INR: International normalized ratio
PMT: Population management tool
QALY: Quality-adjusted life year
UI: Uncertainty interval

## Acknowledgements

None.

## Sources of Funding

This project was supported by grant number R18HS026156 from the Agency for Healthcare Research and Quality. The content is solely the responsibility of the authors and does not necessarily represent the official views of the Agency for Healthcare Research and Quality.

## Disclosures

**KR** has received support from the National Heart, Lung, and Blood Institute (R01HL142834) and CSL Behring, LLC.

**JA** is supported by grant funding from the National Heart, Lung, and Blood Institute (R01HL142834) and AstraZeneca and Bayer.

**DMW** is supported by grant funding from the Agency for Healthcare Research and Quality (R18 HS27960)

**PMH** is supported by grants from the National Heart, Lung, and Blood Institute, the Veterans Affairs HSR&D, and University of Colorado School of Medicine. He had a research agreement with Bristol-Myers Squibb through the University of Colorado focused on atrial fibrillation. He serves as the Deputy Editor for *Circulation: Cardiovascular Quality and Outcomes*.

**JBK** is supported by grant funding from the Agency for Healthcare Research and Quality; the National Heart, Lung, and Blood Institute; and the National Institute on Aging.

**BKB** was supported by National Heart, Lung, and Blood Institute (K01HL140170).

**All other authors** have no relationships to disclose.

## Notes

### Funding Statement

This work was funded by a grant (R18HS026156) from the Agency for Healthcare Research and Quality (AHRQ).

### Author Declarations

Kaiser Permanente's interregional Institutional Review Board (KPiIRB)

## REFERENCES

1. Birger M, Kaldjian AS, Roth GA, Moran AE, Dieleman JL, Bellows BK. Spending on Cardiovascular Disease and Cardiovascular Risk Factors in the United States: 1996 to 2016. Circulation. 2021;144:271–282. doi: 10.1161/CIRCULATIONAHA.120.053216

2. Troy A, Anderson TS. National Trends in Use of and Spending on Oral Anticoagulants Among US Medicare Beneficiaries From 2011 to 2019. JAMA Health Forum. 2021;2:e211693–e211693. doi: 10.1001/jamahealthforum.2021.1693

3. Ko D, Lin KJ, Bessette LG, Lee SB, Walkey AJ, Cheng S, Kim E, Glynn RJ, Kim DH. Trends in Use of Oral Anticoagulants in Older Adults With Newly Diagnosed Atrial Fibrillation, 2010-2020. JAMA Network Open. 2022;5:e2242964–e2242964. doi: 10.1001/jamanetworkopen.2022.42964

4. Navar AM, Kolkailah AA, Overton R, Shah NP, Rousseau JF, Flaker GC, Pignone MP, Peterson ED. Trends in Oral Anticoagulant Use Among 436 864 Patients With Atrial Fibrillation in Community Practice, 2011 to 2020. J Am Heart Assoc. 2022;11:e026723. doi: 10.1161/jaha.122.026723

5. Burnett AE, Barnes GD. A call to action for anticoagulation stewardship. Res Pract Thromb Haemost. 2022;6:e12757. doi: 10.1002/rth2.12757

6. Derington CG, Goodrich GK, Xu S, Clark NP, Reynolds K, An J, Witt DM, Smith DH, O’Keeffe-Rosetti M, Lang DT, et al. Association of Direct Oral Anticoagulation Management Strategies With Clinical Outcomes for Adults With Atrial Fibrillation. JAMA Netw Open. 2023;6:e2321971. doi: 10.1001/jamanetworkopen.2023.21971

7. Fang MC, Go AS, Chang Y, Hylek EM, Henault LE, Jensvold NG, Singer DE. Death and disability from warfarin-associated intracranial and extracranial hemorrhages. Am J Med. 2007;120:700–705. doi: 10.1016/j.amjmed.2006.07.034

8. Harrington AR, Armstrong EP, Nolan PE, Jr., Malone DC. Cost-effectiveness of apixaban, dabigatran, rivaroxaban, and warfarin for stroke prevention in atrial fibrillation. Stroke. 2013;44:1676–1681. doi: 10.1161/strokeaha.111.000402

9. Hemphill JC, 3rd, Farrant M, Neill TA, Jr. Prospective validation of the ICH Score for 12-month functional outcome. Neurology. 2009;73:1088–1094. doi: 10.1212/WNL.0b013e3181b8b332

10. Kamel H, Johnston SC, Easton JD, Kim AS. Cost-effectiveness of dabigatran compared with warfarin for stroke prevention in patients with atrial fibrillation and prior stroke or transient ischemic attack. Stroke. 2012;43:881–883. doi: 10.1161/strokeaha.111.641027

11. Lioutas VA, Goyal N, Katsanos AH, Krogias C, Zand R, Sharma VK, Varelas P, Malhotra K, Paciaroni M, Sharaf A, et al. Clinical Outcomes and Neuroimaging Profiles in Nondisabled Patients With Anticoagulant-Related Intracerebral Hemorrhage. Stroke. 2018;49:2309–2316. doi: 10.1161/strokeaha.118.021979

12. Toyoda K, Yoshimura S, Nakai M, Koga M, Sasahara Y, Sonoda K, Kamiyama K, Yazawa Y, Kawada S, Sasaki M, et al. Twenty-Year Change in Severity and Outcome of Ischemic and Hemorrhagic Strokes. JAMA Neurology. 2022;79:61–69. doi: 10.1001/jamaneurol.2021.4346

13. ElHabr AK, Katz JM, Wang J, Bastani M, Martinez G, Gribko M, Hughes DR, Sanelli P. Predicting 90-day modified Rankin Scale score with discharge information in acute ischaemic stroke patients following treatment. BMJ Neurol Open. 2021;3:e000177. doi: 10.1136/bmjno-2021-000177

14. Arias E, Xu JQ. United States life tables, 2019. National Vital Statistics Reports; vol 70 no 19. Hyattsville, MD: National Center for Health Statistics. 2022. DOI: 10.15620/cdc:113096.

15. US Department of Veterans Affairs. VA Federal Supply Schedule Service. Updated March 2024. Accessed March 17, 2024.

16. Basu A. Estimating Costs and Valuations of Non-Health Benefits in Cost-Effectiveness Analysis. In: Neumann PJ, Ganiats TG, Russell LB, Sanders GD, Siegel JE, eds. Cost-Effectiveness in Health and Medicine. Oxford University Press; 2016:220–229.

17. Centers for Medicare & Medicaid Services (CMS). Physician Fee Schedule. https://www.cms.gov/medicare/physician-fee-schedule/search.

18. May 2021 National Occupational Employment and Wage Estimates. United States. Occupational Employment and Wage Statistics. U.S. Bureau of Labor Statistics. Accessed October 2022. https://www.bls.gov/oes/current/oes_nat.htm#31-0000.

19. Peterson C, Xu L, Florence C, Grosse SD, Annest JL. Professional Fee Ratios for US Hospital Discharge Data. Med Care. 2015;53:840–849. doi: 10.1097/mlr.0000000000000410

20. HCUP National Inpatient Sample (NIS). Healthcare Cost and Utilization Project (HCUP). 2018. Agency for Healthcare Research and Quality, Rockville, MD. www.hcup-us.ahrq.gov/nisoverview.jsp

21. Medical expenditure panel survey (MEPS). Agency for Healthcare Research and Quality. Available: https://www.meps.ahrq.gov/mepsweb/data_stats/download_data_files.jsp

22. Sullivan PW, Ghushchyan V. Preference-Based EQ-5D index scores for chronic conditions in the United States. Med Decis Making. 2006;26:410–420. doi: 10.1177/0272989x06290495

23. Gage BF, Cardinalli AB, Owens DK. The effect of stroke and stroke prophylaxis with aspirin or warfarin on quality of life. Arch Intern Med. 1996;156:1829–1836.

24. O’Brien CL, Gage BF. Costs and Effectiveness of Ximelagatran for Stroke Prophylaxis in Chronic Atrial Fibrillation. JAMA. 2005;293:699–706. doi: 10.1001/jama.293.6.699

25. Harrington AR, Armstrong EP, Nolan PE, Malone DC. Cost-Effectiveness of Apixaban, Dabigatran, Rivaroxaban, and Warfarin for Stroke Prevention in Atrial Fibrillation. Stroke. 2013;44:1676–1681. doi: doi:10.1161/STROKEAHA.111.000402

26. Sullivan PW, Arant TW, Ellis SL, Ulrich H. The cost effectiveness of anticoagulation management services for patients with atrial fibrillation and at high risk of stroke in the US. Pharmacoeconomics. 2006;24:1021–1033. doi: 10.2165/00019053-200624100-00009

27. Sanders GD, Neumann PJ, Basu A, Brock DW, Feeny D, Krahn M, Kuntz KM, Meltzer DO, Owens DK, Prosser LA, et al. Recommendations for Conduct, Methodological Practices, and Reporting of Cost-effectiveness Analyses: Second Panel on Cost-Effectiveness in Health and Medicine. JAMA. 2016;316:1093–1103. doi: 10.1001/jama.2016.12195

28. Kazi DS, Abdullah AR, Arnold SV, Basu A, Bellows BK, Breathett K, Chew DS, Cohen DJ, DeJong C, Heidenreich PA, et al. 2025 AHA/ACC Statement on Cost/Value Methodology in Clinical Practice Guidelines (Update From 2014 Statement): A Report of the American College of Cardiology/American Heart Association Joint Committee on Clinical Practice Guidelines. Circulation. 2025;152:e332–e358. doi: 10.1161/cir.0000000000001377

29. Schnabel RB, Yin X, Gona P, Larson MG, Beiser AS, McManus DD, Newton-Cheh C, Lubitz SA, Magnani JW, Ellinor PT, et al. 50 year trends in atrial fibrillation prevalence, incidence, risk factors, and mortality in the Framingham Heart Study: a cohort study. Lancet. 2015;386:154–162. doi: 10.1016/s0140-6736(14)61774-8

30. Huang Y, Chen SH, Wang Y, Zhao B. Cost Effectiveness Of Direct Oral Anticoagulants (DOACS) Compared With Warfarin: A Literature Review. Value in Health. 2018;21:S60. doi: 10.1016/j.jval.2018.04.367

31. Raccah BH, Erlichman Y, Pollak A, Matok I, Muszkat M. Prescribing Errors With Direct Oral Anticoagulants and Their Impact on the Risk of Bleeding in Patients With Atrial Fibrillation. J Cardiovasc Pharmacol Ther. 2021;26:601–610. doi: 10.1177/10742484211019657

32. Armbruster AL, Buehler KS, Min SH, Riley M, Daly MW. Evaluation of dabigatran for appropriateness of use and bleeding events in a community hospital setting. American health & drug benefits. 2014;7:376–384.

33. Alghadeer S, Hornsby L. Assessment of novel oral anticoagulant use within a community teaching hospital. Saudi Pharmaceutical Journal. 2017;25:93–98. doi: 10.1016/j.jsps.2016.02.002

34. Basaran O, Filiz Basaran N, Cekic EG, Altun I, Dogan V, Mert GO, Mert KU, Akin F, Soylu MO, Memic Sancar K, et al. PRescriptiOn PattERns of Oral Anticoagulants in Nonvalvular Atrial Fibrillation (PROPER study). Clinical and applied thrombosis/hemostasis : official journal of the International Academy of Clinical and Applied Thrombosis/Hemostasis. 2015. doi: 10.1177/1076029615614395

35. Ashjian E, Kurtz B, Renner E, Yeshe R, Barnes GD. Evaluation of a pharmacist-led outpatient direct oral anticoagulant service. American journal of health-system pharmacy : AJHP : official journal of the American Society of Health-System Pharmacists. 2017;74:483–489. doi: 10.2146/ajhp151026

36. Steinberg BA, Shrader P, Thomas L, Ansell J, Fonarow GC, Gersh BJ, Kowey PR, Mahaffey KW, Naccarelli G, Reiffel J, et al. Off-Label Dosing of Non-Vitamin K Antagonist Oral Anticoagulants and Adverse Outcomes: The ORBIT-AF II Registry. Journal of the American College of Cardiology. 2016;68:2597–2604. doi: 10.1016/j.jacc.2016.09.966

37. Larock AS, Mullier F, Sennesael AL, Douxfils J, Devalet B, Chatelain C, Dogne JM, Spinewine A. Appropriateness of prescribing dabigatran etexilate and rivaroxaban in patients with nonvalvular atrial fibrillation: a prospective study. The Annals of pharmacotherapy. 2014;48:1258–1268. doi: 10.1177/1060028014540868

38. Al Rowily A, Jalal Z, Price MJ, Abutaleb MH, Almodiaemgh H, Al Ammari M, Paudyal V. Prevalence, contributory factors and severity of medication errors associated with direct-acting oral anticoagulants in adult patients: a systematic review and meta-analysis. Eur J Clin Pharmacol. 2022;78:623–645. doi: 10.1007/s00228-021-03212-y

39. Geller AI, Shehab N, Lovegrove MC, Weidle NJ, Budnitz DS. Bleeding related to oral anticoagulants: Trends in US emergency department visits, 2016-2020. Thrombosis Research. 2023;225:110–115. doi: 10.1016/j.thromres.2023.03.010

40. Budnitz DS, Lovegrove MC, Shehab N, Richards CL. Emergency hospitalizations for adverse drug events in older Americans. N Engl J Med. 2011;365:2002–2012. doi: 10.1056/NEJMsa1103053

41. Budnitz DS, Shehab N, Lovegrove MC, Geller AI, Lind JN, Pollock DA. US Emergency Department Visits Attributed to Medication Harms, 2017-2019. Jama. 2021;326:1299–1309. doi: 10.1001/jama.2021.13844

42. Fang H, Frean M, Sylwestrzak G, Ukert B. Trends in Disenrollment and Reenrollment Within US Commercial Health Insurance Plans, 2006-2018. JAMA Network Open. 2022;5:e220320–e220320. doi: 10.1001/jamanetworkopen.2022.0320

43. Meyers DJ, Belanger E, Joyce N, McHugh J, Rahman M, Mor V. Analysis of Drivers of Disenrollment and Plan Switching Among Medicare Advantage Beneficiaries. JAMA Internal Medicine. 2019;179:524–532. doi: 10.1001/jamainternmed.2018.7639

44. Guo Y, Guo J, Shi X, Yao Y, Sun Y, Xia Y, Yu B, Liu T, Chen Y, Lip GYH, et al. Mobile health technology-supported atrial fibrillation screening and integrated care: A report from the mAFA-II trial Long-term Extension Cohort. Eur J Intern Med. 2020;82:105–111. doi: 10.1016/j.ejim.2020.09.024

45. Allen AL, Lucas J, Parra D, Spoutz P, Kibert JL, 2nd, Ragheb B, Chia L, Sipe A. Shifting the Paradigm: A Population Health Approach to the Management of Direct Oral Anticoagulants. J Am Heart Assoc. 2021;10:e022758. doi: 10.1161/jaha.121.022758

46. Dorsch MP, Chen CS, Allen AL, Sales AE, Seagull FJ, Spoutz P, Sussman JB, Barnes GD. Nationwide Implementation of a Population Management Dashboard for Monitoring Direct Oral Anticoagulants: Insights From the Veterans Affairs Health System. Circulation: Cardiovascular Quality and Outcomes. 2023;16:e009256. doi: 10.1161/CIRCOUTCOMES.122.009256

47. Barnes GD, Chen C, Holleman R, Errickson J, Seagull FJ, Dorsch MP, Allen AL, Spoutz P, Sussman JB. Pharmacist Use of a Population Management Dashboard for Safe Anticoagulant Prescribing: Evaluation of a Nationwide Implementation Effort. Journal of the American Heart Association. 2024;13:e035859. doi: 10.1161/JAHA.124.035859

48. Joglar JA, Chung MK, Armbruster AL, Benjamin EJ, Chyou JY, Cronin EM, Deswal A, Eckhardt LL, Goldberger ZD, Gopinathannair R, et al. 2023 ACC/AHA/ACCP/HRS Guideline for the Diagnosis and Management of Atrial Fibrillation: A Report of the American College of Cardiology/American Heart Association Joint Committee on Clinical Practice Guidelines. Circulation. 2024;149:e1–e156. doi: 10.1161/CIR.0000000000001193

49. Joosten LPT, van Doorn S, van de Ven PM, Köhlen BTG, Nierman MC, Koek HL, Hemels MEW, Huisman MV, Kruip M, Faber LM, et al. Safety of Switching From a Vitamin K Antagonist to a Non–Vitamin K Antagonist Oral Anticoagulant in Frail Older Patients With Atrial Fibrillation: Results of the FRAIL-AF Randomized Controlled Trial. Circulation. 2024;149:279–289. doi: 10.1161/CIRCULATIONAHA.123.066485

50. Holbrook A, Schulman S, Witt DM, Vandvik PO, Fish J, Kovacs MJ, Svensson PJ, Veenstra DL, Crowther M, Guyatt GH. Evidence-based management of anticoagulant therapy: Antithrombotic Therapy and Prevention of Thrombosis, 9th ed: American College of Chest Physicians Evidence-Based Clinical Practice Guidelines. Chest. 2012;141:e152S–e184S. doi: 10.1378/chest.11-2295

51. Jones AE, King JB, Kim K, Witt DM. The role of clinical pharmacy anticoagulation services in direct oral anticoagulant monitoring. J Thromb Thrombolysis. 2020;50:739–745. doi: 10.1007/s11239-020-02064-0

52. O’Brien CL, Gage BF. Costs and effectiveness of ximelagatran for stroke prophylaxis in chronic atrial fibrillation. Jama. 2005;293:699–706. doi: 10.1001/jama.293.6.699

